# Automated Echocardiographic Detection of Mitral Valve Prolapse and Mitral Regurgitation with Video-based Artificial Intelligence Algorithms

**DOI:** 10.64898/2026.02.26.26347229

**Authors:** Minhaj U. Ansari, Joshua P. Barrios, Lionel Tastet, Rohit Jhawar, Luca Cristin, Amy Rich, Dwight Bibby, Qizhi Fang, Farzin Arya, Valentina Crudo, Thuy Nguyen, Dipan J. Shah, Francesca N. Delling, Geoffrey H. Tison

## Abstract

**Aims:** We aimed to develop and evaluate fully automated artificial intelligence (AI) system. for detection of mitral valve prolapse (MVP) and mitral regurgitation (MR) from echocardiographic studies.

**Methods and Results:** We used a dataset of 24,869 echocardiographic studies from the University of California San Francisco (UCSF) to train a multi-view deep neural network (DNN) to detect MVP using apical 4-chamber, 2-chamber, and parasternal long-axis views. A separate dataset of 27,906 studies from UCSF was used to train a second multi-view DNN model to detect moderate-to-severe or severe MR using color Doppler in the same views. External validation was performed on echocardiographic MVP videos from Houston Methodist Hospital.

The DNN model for MVP detection achieved an AUC of 0.917 (95% CI: 0.899-0.934), with stronger performance in those with mitral annular disjunction or bileaflet MVP. External validation for MVP detection in a geographically and demographically distinct population yielded an AUC of 0.835 (95% CI: 0.803-0.869). The DNN for detection of moderate-to-severe or severe MR in patients with concurrent MVP achieved an AUC of 0.877 (95% CI: (0.805-0.939).

**Conclusions:** AI algorithms can perform automatic detection of MVP and clinically significant MR from echocardiogram studies with high performance. The MVP DNN performed particularly well for more severe MVP phenotypes such as mitral annular disjunction or bileaflet MVP. These algorithms could provide a novel approach for automated, accurate, and rapid diagnosis of MVP and its common clinical sequelae across institutions.

**Graphical Abstract:** 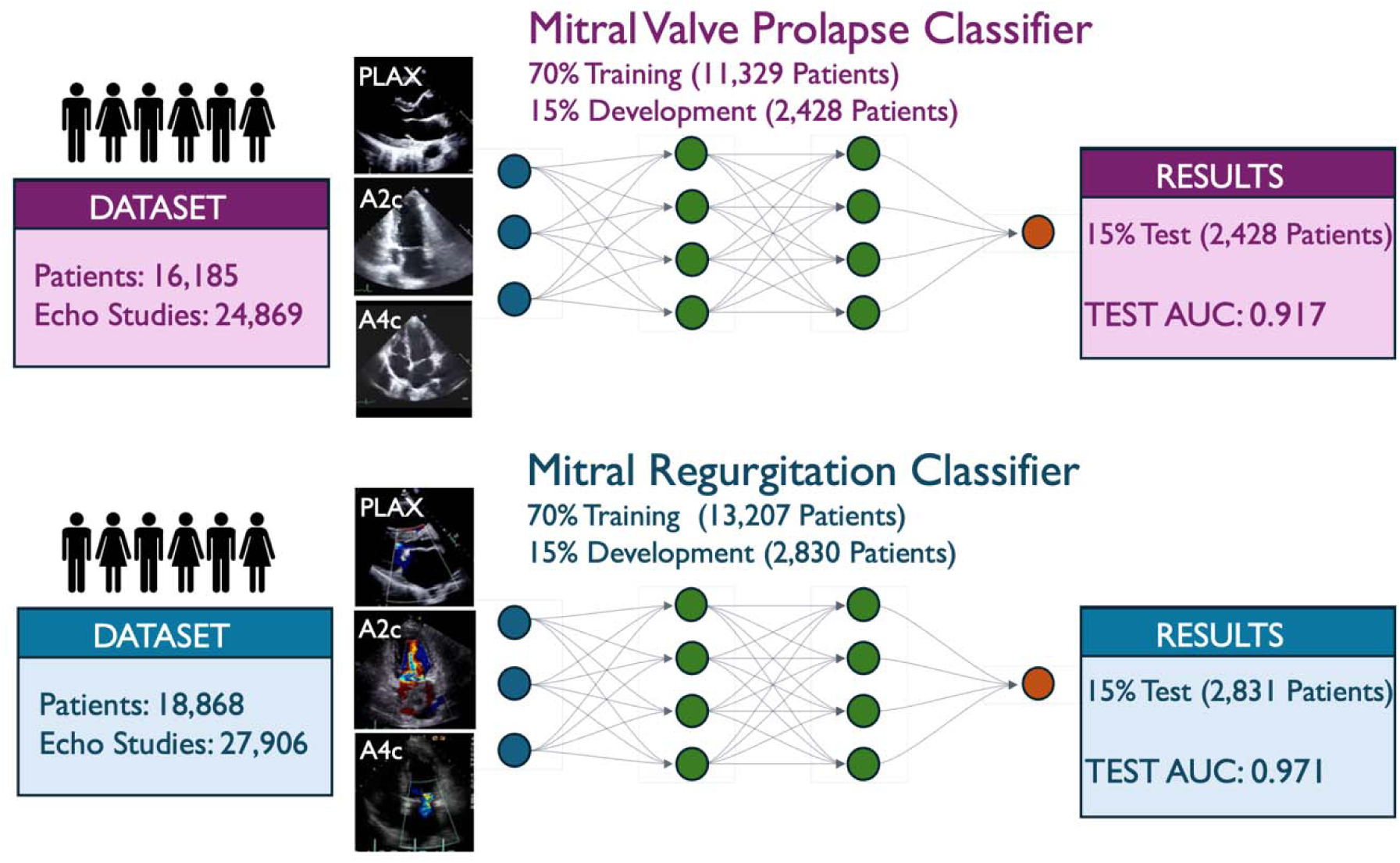

## Introduction

Mitral valve prolapse (MVP) is a common valvular disease affecting 2% to 3% of individuals in the general population, and is a leading cause of mitral regurgitation (MR) requiring surgical repair or replacement(1–3). In the absence of significant MR, MVP was previously considered a benign condition. However, it is increasingly recognized that a subset of individuals with MVP has increased risk of major clinical complications (~1% to 2% yearly) including atrial or ventricular arrhythmias, cardiomyopathy, congestive heart failure, sudden arrhythmic death, and excess mortality(4–9). Therefore, early detection and accurate assessment of MVP are critical for optimal risk stratification, clinical monitoring, and timely intervention.

Echocardiography remains the first-line imaging modality for diagnosis of MVP. It allows for assessment of morpho-functional abnormalities of the mitral valve and related structures including presence of mitral annular disjunction (MAD), MR severity, and structural and functional consequences on the myocardium(10). However, accurate diagnosis of MVP can be challenging, especially due the saddle shaped and dynamic nature of the mitral valve leaflets. Such complexity often leads to inter-observer variability and missed MVP diagnosis, even among experienced echocardiographic readers. Such variability and underdiagnosis may be even more common in under-resourced settings, making it possible that more standardized and objective tools may be needed to improve the diagnosis of MVP and its sequelae.

In recent years, artificial intelligence (AI) algorithms have emerged as promising tools for screening, diagnosis, and image interpretation across a range of cardiovascular conditions(11–13). While some work has begun to examine how AI may assist for valve pathologies(13,14), MVP-specific efforts have been limited and relied on small cohorts(15,16). We aimed to employ a novel video-based deep neural network (DNN) architecture to automatically detect MVP from echocardiographic videos. Furthermore, because MVP encompasses abnormalities in both structure and movement we employed a novel video-based DNN architecture that considers multiple echo views simultaneously, instead of relying only on a single echo video or still-frame echo images. Finally, because MVP patients are at greater risk of MR due to leaflet malcoaptation from MVP, we also aimed to evaluate whether DNNs could help to detect MR in MVP patients.

## Methods

### Study Participants and Study Datasets

For this cross-sectional study, we obtained transthoracic echocardiograms of adult patients (≥18 years old) performed at the University of California San Francisco (UCSF) between January 1, 2000, and December 31, 2023. Stress echocardiograms and transesophageal echocardiograms were excluded from analysis.

#### MVP Dataset

The UCSF MVP registry of 718 MVP patients was constructed by chart review and stored in an electronic database(5). All echocardiograms in the UCSF MVP registry were re-analyzed for MVP according to a systematic MVP protocol by MVP expert Dr. Francesca Delling (Co-senior author) and 2 veteran research sonographers (co-authors FA and DB), constituting a gold-standard, research-grade MVP adjudication that is substantially more reliable than the clinical echo report alone. After excluding patients <18 years old or without a UCSF echocardiogram, all echocardiograms were obtained for 661 MVP patients (**Figure 1**). We further excluded echocardiograms performed either before the date of MVP diagnosis or after mitral valve repair. Moreover, we excluded echocardiograms which did not contain at least one view from our three views of interest: -apical 2-chamber (A2C), 4-chamber (A4C), and parasternal long axis (PLAX)-leading to exclusion of 8 patients. We used PLAX rather than apical 3-chamber views, as both provide similar long-axis visualization of A2/P2 scallops, and inclusion of 2- and 4-chamber views enabled assessment of additional scallops not seen in long-axis views. These exclusion criteria resulted in a final MVP cohort consisting of 584 patients and a total of 1,492 echocardiograms.

**Figure 1:**
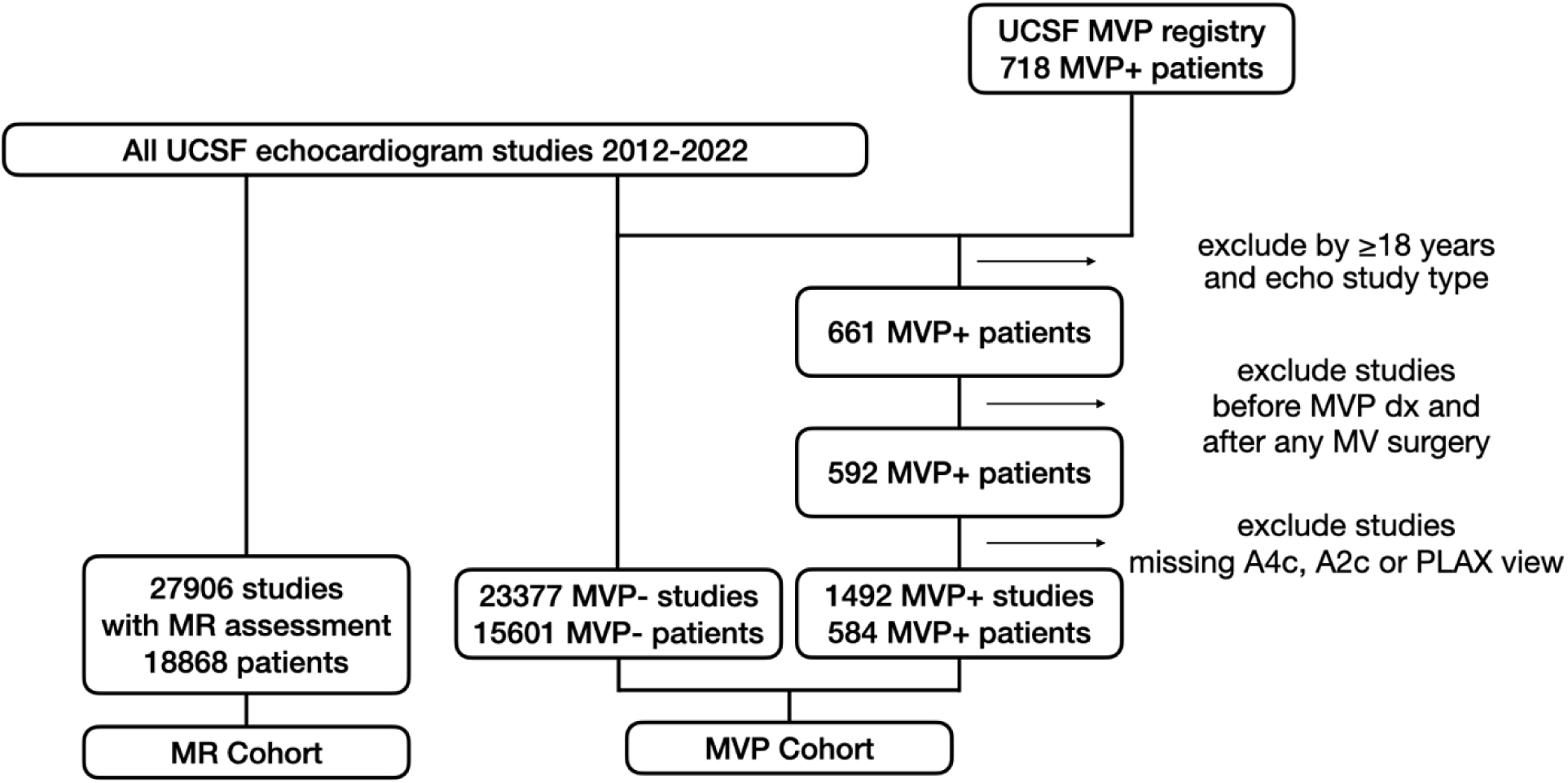
Data Creation Flowchart

**Figure 2:**
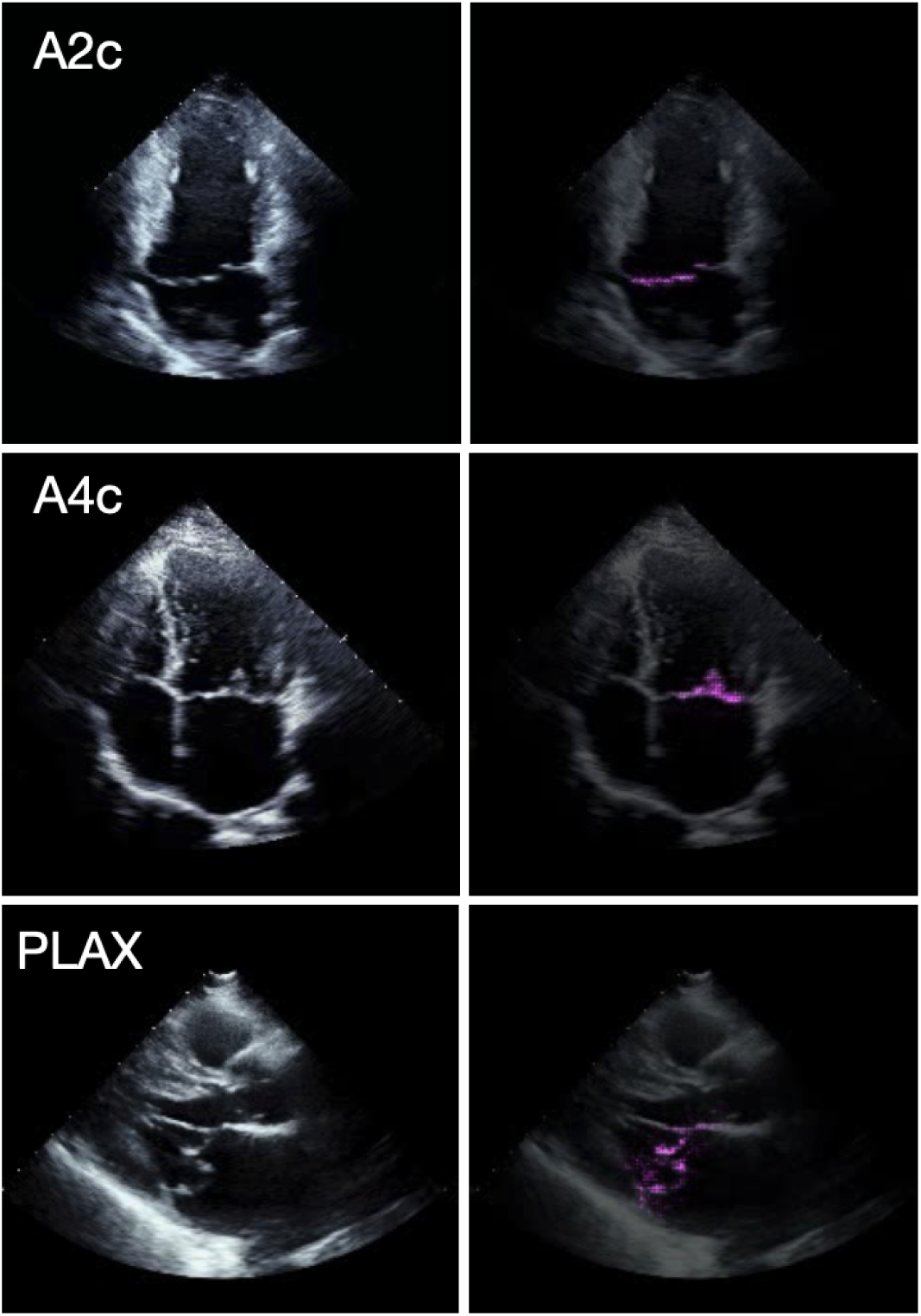
Echocardiogram regions of greater importance for mitral valve prolapse deep neural network Left: Frames taken from echocardiographic videos of mitral valve prolapse (MVP) patients from the three views used in our study: apical 2-chamber (A2c), apical 4-chamber (A4c), and parasternal long-axis (PLAX). Right: The same frames are overlayed with the pixel-wise Grad-CAM heatmaps. Magenta pixels highlight regions of the image that most strongly contributed to the model’s prediction of MVP.

#### Control Dataset

To accommodate our data augmentation approach, described below, we constructed a control cohort of non-MVP control studies randomly sampled from echocardiograms performed at UCSF during the same period. This cohort was age- and sex-matched to the MVP cases. From these, similarly to the MVP sample, we excluded stress/transesophageal studies and studies that did not contain at least one clip from all three of our views of interest. Our non-MVP control dataset consisted of 23,377 studies from 15,601 patients.

#### External Validation Dataset

For external validation of our MVP DNN model, we obtained 118 MVP studies from 118 patients acquired at the Houston Methodist DeBakey Heart & Vascular Center (Houston, TX). We constructed a non-MVP control cohort for this validation using 4,068 random UCSF echocardiograms from 3,728 non-MVP patients that were not in the UCSF training or development sets. Because this external dataset combines MVP cases from Houston with non-MVP controls from UCSF, it represents a “hybrid” external validation set.

#### MR Detection Dataset

Given that MR is a common sequela of MVP (1,22), we collected a total of 27,906 echocardiographic studies from 18,868 patients, including 537 MR-positive and 27,369 MR-negative studies (**Figure 1**).

### Echocardiography

Transthoracic echocardiograms were performed using commercially available ultrasound systems. MVP and MAD labels were derived from the UCSF MVP registry, the echocardiograms for which were re-adjudicated for MVP and MAD by an experienced echocardiographer and two senior research sonographers. MVP was defined as systolic displacement of one or both mitral leaflets ≥2 mm beyond the mitral annulus in a parasternal or apical 3-chamber long-axis view(17,18) MAD was defined as a separation between the basal inferolateral LV myocardium and the LA/mitral leaflet junction and was assessed in parasternal long-axis view, as previously described(5). The MVP phenotype was further classified as bileaflet versus single leaflet prolapse. MR severity was assessed using a multi-parameter approach as recommended by guidelines(10). MR severity was graded for each echocardiogram as follows: no/trace, mild, mild-moderate, moderate, moderate-severe or severe. All MR labels were derived at the echo study level from complete clinical report, reflecting cardiologist assessment based on all available views and doppler imaging. For the MR DNN classifier, we defined “MR+” as moderate-severe or severe MR, and MR- as equal or less than moderate MR.(5)

### Model design and training

All model design and training was done in Python 3.8.8 using the Pytorch 1.8 library(19). We tested the X3D-M 3-D convolutional video architecture (20) using each of our three chosen echo views independently, along with a custom multi-view implementation of X3D-M that incorporates information from all three views into a prediction. This method concatenates the 3-D embeddings of each view along a fourth “view” dimension. The resulting tensor is reshaped and flattened across the spatiotemporal dimensions, allowing a bespoke 3-D convolutional block to fuse information across views before passing the multi-view embedding through subsequent classification layers. This multi-view method was superior to single-view X3D models using individual views; and was therefore used for all main analyses (Supplemental Table 1).

All DNN models used a binary classification using a sigmoid decision head with the binary cross entropy loss function. For training, datasets were divided into training, development, and test sets at a 70:15:15 ratio by patient. Performance on the development sets was used for evaluation during training and for choosing the best checkpoint. All reported metrics are on the held-out test sets. All decision thresholds were chosen to maximize the geometric mean of sensitivity and specificity. To train the MVP DNN, we performed hyperparameter sweeps over learning rate, and the threshold and patience values of the Pytorch ReduceLROnPlateau learning rate scheduler. To train the MR DNN, we used the LRFinder algorithm (21) to find the best LR and then used the Pytorch CosineAnnealingWarmRestarts learning rate scheduler. All models were trained for 50 epochs, and the best checkpoint was chosen based on the AUC of the validation set.

### Data processing

Pixel data were extracted from the Digital Imaging and Communications in Medicine (DICOM) format, masked to remove any information outside the scanning sector (e.g., annotations), square cropped, scaled to 224×224 pixels, and then saved in AVI (Audio Video Interleave) format. DNN input consisted of 32 frames of the echocardiogram videos, temporally subsampled by a factor of 2. Echocardiogram video frame rates were 33±17 frames per second. All echocardiogram studies, including external MVP studies from Houston Methodist DeBakey, were analyzed with a previously trained view classifier DNN. The classifier automatically identified clips corresponding to the three predefined mitral valve views selected for this study based on clinical relevance (A2C, A4C, PLAX) and detected the presence of color Doppler in each clip. For MVP modeling, the pipeline automatically selected non–color Doppler clips from these views. For MR modeling, the pipeline automatically selected color Doppler clips from the same views. This clip selection strategy enabled fully automated end-to-end analysis of transthoracic echocardiogram studies. We used data augmentation techniques to enhance model robustness. Images were randomly cropped and resized to 224 pixels, with scales ranging from 95% to 100% of the original image size and aspect ratios varying between 0.95 and 1.05. Additional augmentations included color jitter with brightness, contrast, and saturation values randomly adjusted between 80% and 120% of their original levels, with no hue variation, and random rotations between −5 and 5 degrees.

Because our multi-view DNN required an input of 3 views taken from the same echocardiographic study, and because there was often more than one example of a view obtained during a study, we employed an additional data augmentation strategy during training only for the MVP DNN models. In the training dataset, we randomly selected one video from each required view (A4C, A2C, PLAX) if more than one was available for a given study, creating multiple sets of 3 views per study. This was done for all MVP cases and non-MVP controls. During training, we used all available 3-view sets from MVP cases, while preferentially taking 3-view sets from separate non-MVP control patients to increase the individual variation within the controls from which the DNN was able to learn. For evaluation in the MVP test-dataset, only one set of views was selected per study. To train the MR DNN, we used the same three views (A4C, A2C, PLAX) that contained color Doppler windows and the same data augmentation strategy as described for the MVP model. For evaluation in the MR test dataset, only one set of 3 color Doppler views was selected per echocardiogram.

### Consent and Ethical approval

This study was reviewed and received approval from the UCSF Institutional Review Board and the Houston Methodist Review Board; at both sites, the need to obtain patient consent was waived as the study used retrospective data.

## Results

### Automated detection of MVP

The mean age of the UCSF MVP cohort was 61±16 years and 50% were male (**Table 1**). Among MVP patients, 46% had bileaflet MVP, and 42% had MAD. The MVP multi-view DNN demonstrated strong performance, achieving an AUC of 0.917 (95% CI: 0.899-0.934) for detecting MVP, with a sensitivity 0.797 (95% CI: 0.754-0.841) and a specificity 0.893 (95% CI: 0.884-0.901; **Table 2**). Notably, this multi-view MVP DNN had higher discrimination than any DNN trained using each of the three single-view models individually to detect MVP (PLAX view: 0.879; A4C view: 0.862, and A2C view: 0.862, **Supplemental Table 1**). When examined according to age, the multi-view DNN showed higher discrimination for patients ≤62 years of age (i.e, median age of the MVP dataset), with an AUC of 0.950 (95% CI: 0.933-0.966) compared to an AUC of 0.880 (95% CI: 0.848-0.909) for patients >62 years of age. Sensitivity and specificity followed a similar pattern (**Table 2**). MVP DNN performance was comparable between male and female patients (**Table 2**).

**Table 1:** Characteristics of the MVP Datasets.

| Datasets | UCSF Training Dataset | UCSF Development Dataset | UCSF Test Dataset | Houston Methodist Hybrid-External Validation Dataset |
| --- | --- | --- | --- | --- |
| **Clinical characteristics** |  |  |  |  |
| **Echo Studies** | 17449 | 3698 | 3722 | 4186 |
| **Patients** | 11329 | 2428 | 2428 | 3846 |
| **Male sex** | 5671 (50%) | 1226 (50%) | 1217 (50%) | 1956 (51%) |
| **Age (mean, std)** | 61 (16) | 61 (17) | 61 (16) | 61 (18) |
| **Body surface area, m<sup>2</sup> (mean, std)** | 1.86 (0.27) | 1.85 (0.27) | 1.86 (0.27) | 1.86 (0.28) |
| **MVP Patients** | 414 | 89 | 81 | 118 |
| **Bileaflet** | 197 (48%) | 42 (47%) | 31 (38%) | 74 (63%) |
| **MAD** | 173 (42%) | 42 (47%) | 30 (37%) | 1 (1%) |
| **MR+ (Studies)** | 172 (16%) | 23 (10%) | 39 (18%) | 5 (4%) |

Abbreviations: MVP-mitral valve prolapse; MAD-mitral annular disjunction; MR-mitral regurgitation

**Table 2:**
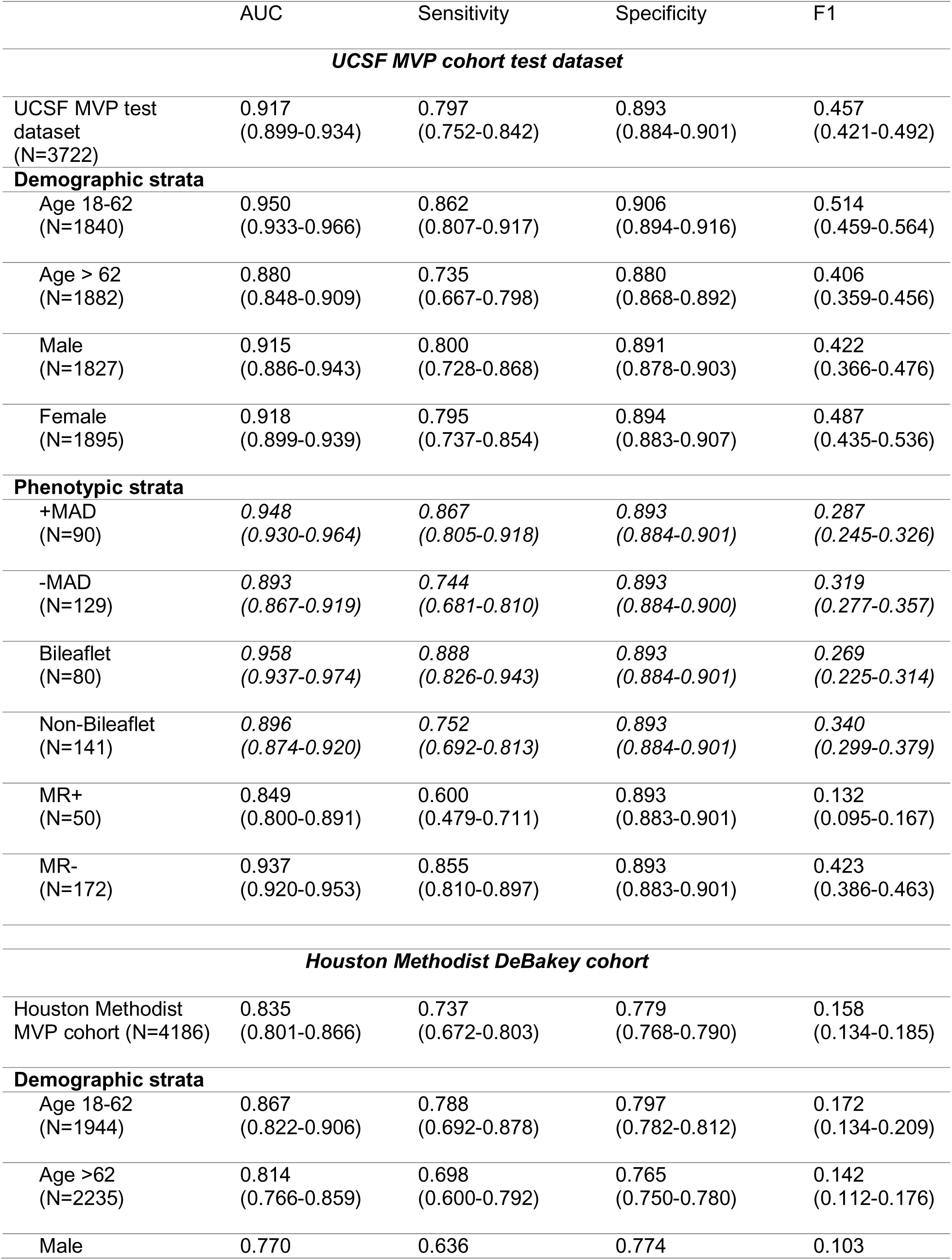
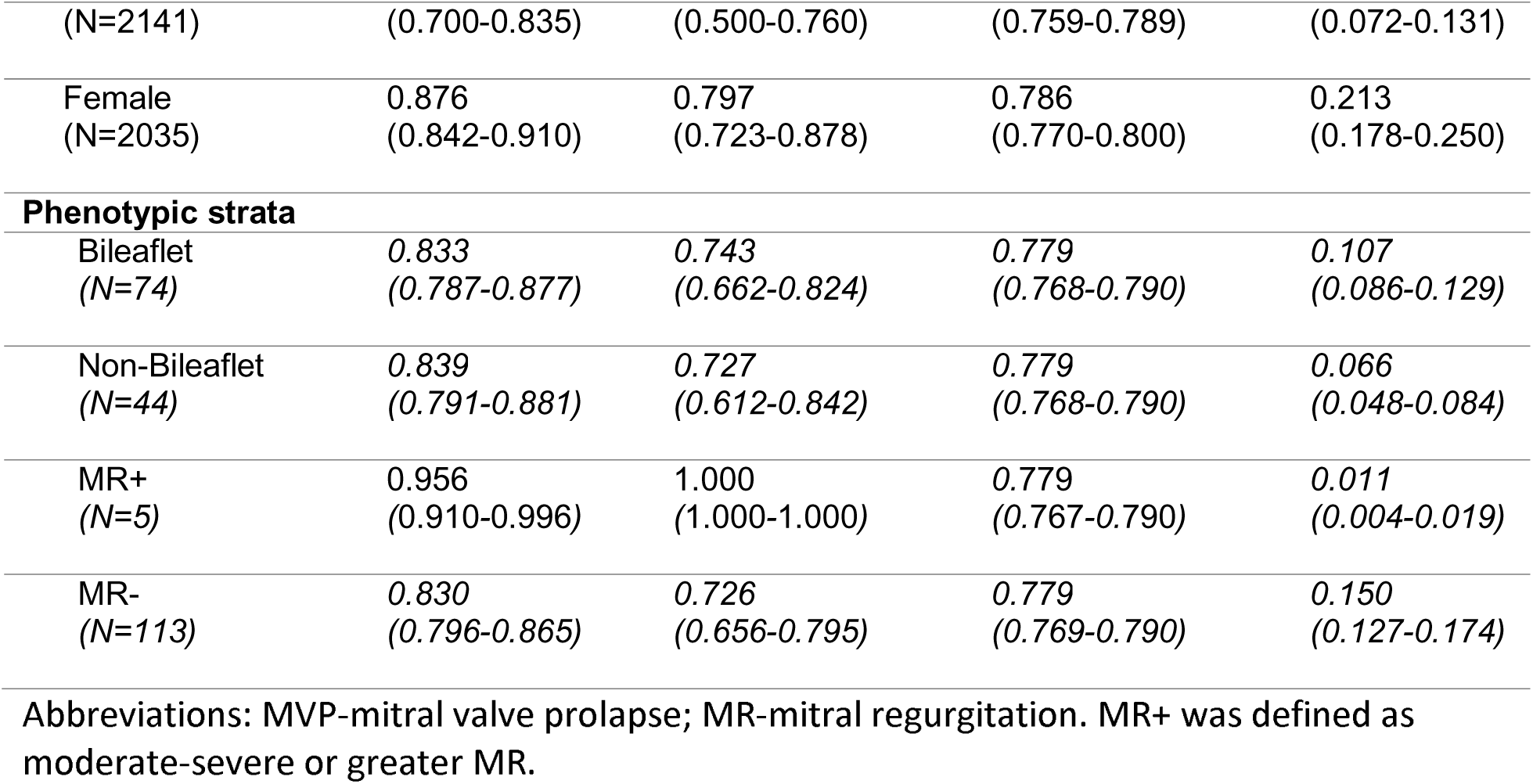
MVP multi-view DNN performance.

We examined the performance of the DNN to detect MVP in the presence of MAD, MR, and bileaflet MVP. Among MVP patients with MAD, the DNN demonstrated excellent discrimination with an AUC of 0.948 (95% CI: 0.930-0.964), exceeding the AUC for the full MVP cohort. Similarly, in patients with bileaflet MVP, the DNN achieved an AUC of 0.958 (95% CI: 0.937-0.974) to detect MVP, also higher than that of the overall MVP cohort. For patients with moderate-severe or greater MR (MR+), the AUC was 0.849 (95% CI 0.805-0.891), slightly lower than the AUC for the overall MVP cohort and in those without moderate-severe or greater MR (**Table 2**).

To evaluate the generalizability of our MVP multi-view DNN, we tested it on external MVP patients obtained from Houston Methodist DeBakey Heart & Vascular center, a geographically and demographically distinct population. In this external validation dataset, the MVP DNN achieved an AUC of 0.835 (95% CI: 0.803-0.869), with a sensitivity of 0.737 (95% CI: 0.672-0.803) and a specificity of 0.779 (95% CI: 0.768-0.789) (**Table 2**). In the Houston Methodist DeBakey dataset, the MVP DNN performed similarly across age strata in the external dataset and performed better in females than males. The performance was similar regardless of bileaflet MVP anatomy and appeared to be higher in those with concurrent moderate-severe or greater MR, although the sample size was small in this sub-stratum. F1 scores are also reported in Table 2, though it should be noted that prevalence differences in the target population (such as between substrata) will impact the F1 score.

We used the Grad-CAM technique to highlight cardiac structures in the echocardiogram that were most strongly associated with the MVP DNN’s predictions. When applied to each of the 3 echocardiographic views, Grad-CAM showed that the MVP DNN consistently focused on the mitral valve region, which would suggest that the DNN uses features of the mitral valve rather than non-valvular features (i.e. myocardium or chamber sizes) to discriminate MVP.

### Automated detection of MR

Given that MR is a common sequela of MVP (1,22), we trained a separate DNN model to detect moderate-severe or severe MR (MR+) from color Doppler echocardiographic videos. A total of 27,906 echocardiographic studies from 18,868 patients were included, with 537 MR+ and 27,369 MR- studies (**Figure 1**). Among the 537 MR+ echocardiographic studies, 381 were in the training dataset (moderate-severe: 235; severe: 146), 75 were in the development dataset (moderate-severe: 49; severe: 26), and 81 were in the test dataset (moderate-severe: 57; severe: 24). The mean patient age was 61±17 years, with approximately 51% of the cohort being male (**Table 3**).

**Table 3:** Characteristics of the MR UCSF Dataset.

|  | Training dataset | Development dataset | Test dataset |
| --- | --- | --- | --- |
| Clinical characteristics |  |  |  |
| Patients | 13207 | 2830 | 2831 |
| Male sex | 6668 (50%) | 1469 (52%) | 1424 (50%) |
| Age (mean, std) | 61 (17) | 60 (17) | 60 (18) |
| MVP | 342 (3%) | 78 (3%) | 73 (3%) |
| Echo Studies | 19588 | 4201 | 4117 |
| MR- |  |  |  |
| None | 4674 (24%) | 1033 (25%) | 982 (24%) |
| Trace | 9229 (47%) | 1897 (45%) | 1963 (48%) |
| Mild | 3845 (20%) | 834 (20%) | 752 (18%) |
| Mild-Moderate | 897 (5%) | 205 (5%) | 206 (5%) |
| Moderate | 562 (3%) | 157 (4%) | 133 (3%) |
| MR+ |  |  |  |
| Moderate-Severe | 235 (1%) | 49 (1%) | 57 (1%) |
| Severe | 146 (1%) | 26 (1%) | 24 (1%) |
| MVP+ |  |  |  |
|  | 761 (4%) | 182 (4%) | 154 (4%) |

Abbreviations: MVP-mitral valve prolapse; MR-mitral regurgitation

The DNN trained to detect MR+ achieved an overall AUC of 0.971 (95% CI: 0.960-0.981), with a sensitivity of 0.877 (95% CI: 0.815-0.938) and a specificity of 0.906 (95%CI 0.898-0.913). The performance was consistent across age strata, dichotomized at the median age of 63 years, and between sexes (**Table 4**). We also examined the performance of the MR DNN to detect MR+ in the presence or absence of MVP. While the AUC was slightly lower to detect moderate-severe or severe MR in patients with MVP (AUC 0.877 95% CI: 0.805-0.939) compared to those without MVP (AUC 0.978 95% CI: 0.968-0.987), discrimination remained robust, even in those with MVP-associated irregular or eccentric MR jets (**Table 4**). In patients with MVP, the MR+ DNN sensitivity remained high but the specificity was lower.

**Table 4:** Mitral regurgitation DNN performance.

|  | AUC | Sensitivity | Specificity | F1 |
| --- | --- | --- | --- | --- |
| MR cohort<br>(N=4,117) | 0.971<br>(0.960-0.981) | 0.877<br>(0.815-0.938) | 0.906<br>(0.898-0.913) | 0.266<br>(0.226-0.308) |
| **Demographic strata** |  |  |  |  |
| Age 18-63<br>(N=2,113) | 0.976<br>(0.962-0.988) | 0.865<br>(0.765-0.950) | 0.934<br>(0.924-0.943) | 0.309<br>(0.239-0.380) |
| Age > 63<br>(N=2,004) | 0.967<br>(0.950-0.981) | 0.886<br>(0.804-0.957) | 0.876<br>(0.864-0.889) | 0.239<br>(0.186-0.291) |
| Male<br>(N=2,075) | 0.963<br>(0.948-0.977) | 0.867<br>(0.782-0.943) | 0.896<br>(0.885-0.907) | 0.264<br>(0.207-0.317) |
| Female<br>(N=2,038) | 0.979<br>(0.965-0.990) | 0.889<br>(0.789-0.969) | 0.916<br>(0.905-0.926) | 0.270<br>(0.204-0.335) |
| **Phenotypic strata** |  |  |  |  |
| Non-MVP<br>(N=3,963) | 0.978<br>(0.968-0.987) | 0.893<br>(0.823-0.959) | 0.914<br>(0.907-0.921) | 0.226<br>(0.186-0.271) |
| MVP<br>(N=89) | 0.877<br>(0.805-0.939) | 0.833<br>(0.667-0.952) | 0.690<br>(0.600-0.783) | 0.545<br>(0.400-0.667) |

Abbreviations: MVP-mitral valve prolapse; MR-mitral regurgitation

## Discussion

In this study, we demonstrated that a multi-view MVP DNN model accurately classifies MVP from complete transthoracic echocardiogram studies with an AUC of 0.917. The MVP DNN performed well in both sex strata and had slightly better performance in younger patients 18-62 years of age. We also showed that a separate MR DNN model can accurately identify clinically significant MR (moderate-severe or severe MR) in MVP patients with an AUC of 0.887, identifying one of the clinically important consequences of MVP. These findings highlight the ability for AI models to assist in the accurate diagnosis of MVP while also identifying MR, its most common clinical consequence, in an automated manner, potentially enabling earlier detection, automated follow-up and personalized patient management.

MVP is a relatively common valvular heart disease that is increasingly recognized to be associated with adverse clinical sequelae including progressive MR, stroke, atrial fibrillation and heart failure(23,24). Patients with MVP have also been shown to have increased risk for ventricular arrhythmias, including the most severe outcome of sudden cardiac death(4,6). Though MVP can be identified through family history and directed screening many MVP cases are incidentally found on routine echocardiograms ordered for other indications. The diagnosis of MVP may be more frequently missed in non-tertiary care, non-surgical centers with rare exposure to valve disease, or when MR is trivial or absent. Under-recognition of MVP may also occur when systolic displacement of mitral valve leaflets is not present in multiple long-axis views when the echocardiographic study lacks systematic interrogation of all leaflet scallops. Even when MVP is diagnosed from transthoracic echocardiograms, distinction between bileaflet and mono-leaflet MVP may be problematic. This may limit overall prognostic classification of MVP, because bileaflet involvement has been shown to be associated with sudden cardiac death in multiple studies(4,6).

Considering the adverse prognosis in a subset of MVP patients, our MVP DNN provides an opportunity to improve the accuracy of MVP diagnosis and to decrease the frequency of missed incidental diagnosis of MVP. This may be particularly beneficial in the common clinical setting where echocardiographic studies are initially obtained for other indications. Use of an MVP DNN to supplement echo interpretation may allow for more standardized and accurate automated detection of MVP across institutions regardless of different levels of reader expertise or disease frequency. Our MVP DNN performed well in the presence or absence of concurrent moderate-severe or severe MR (AUC 0.849 vs AUC of 0.917 in the full MVP cohort), demonstrating that the DNN can identify MVP independently of MR severity. We did observe slightly lower performance of the MVP DNN in patients >62 years old. This may be related to the higher frequency of non-MVP mitral valve pathology in this demographic due to conditions such as annular calcification and leaflet fibrosis which may obscure classical MVP features on echocardiography and potentially confound the model’s predictions. Importantly, many incidentally discovered MVP cases on echocardiography occur in patients under 60 years of age. It is notable that our MVP DNN model demonstrated a particularly strong performance for more advanced forms of myxomatous valve degeneration such as bileaflet MVP and MVP with MAD, both of which may also be more likely to be associated with worse MVP sequalae such as progressive MR, associated cardiomyopathy or ventricular ectopy.Our work contributes to prior efforts that have explored using AI to evaluate the mitral valve, using the largest dedicated cohort of MVP patients. Wifstad et al. described an AI segmentation model that automatically identified segments of the mitral valve that were used to calculate of quantitative valve-related biomarkers of morphology and motion(15). They then examined how these biomarkers differed in three patients with MVP versus other patients without MVP, suggesting at a small-scale that an AI segmentation-based approach could also be used to identify mitral valve anatomy with MVP. Vafaeezadeh et al. used image-based DNNs to classify various MV diseases in a larger cohort including rheumatic mitral stenosis and MVP, though did not report MVP-specific results making it difficult to compare with our results(16). In addition to using a larger cohort dedicated to MVP patients, our work employs a video-based DNN model instead of an image-based model, which analyzes the entire cardiac cycle and therefore removes the need to separately identify the end-systolic phase frame as done by Vafaeezadeh et al.(16). Our work provides an approach to automatically identify MVP that is robust to MV-related structural abnormalities. Our study cohort is additionally distinguished by thorough re-adjudication of MVP with a rigorous protocol, providing precise mitral valve phenotyping.

Although some individuals with MVP have mild or no mitral regurgitation, MVP is clearly also associated with MR and is well known to cause highly-eccentric MR jets directly resulting from the anatomy of the affected leaflet(s). Leaflet displacement in MVP is responsible for poor coaptation of the mitral valve leaflets and consequent MR. Longitudinal studies have shown that MVP can progress to significant MR in a quarter of individuals over a period of 3 to 16 years(25). Furthermore, because MVP-associated MR can often be highly eccentric, MR severity can sometimes be underestimated by less-experienced cardiologists leading to delayed surgical referrals. Because of the close interplay between MVP and MR, we trained a second MR DNN model to detect moderate-severe or greater MR. Our results demonstrate that our MR DNN performs quite well when detecting moderate-severe or greater MR even in MVP patients, achieving an AUC of 0.877. If deployed alongside our MVP DNN, the MR DNN would serve to identify more clinically significant MR amongst patients identified to have MVP, identifying candidates at greater risk of MR progression, more likely to benefit from mitral valve surgery and possibly increased clinical consequences of MVP, such as atrial enlargement or atrial fibrillation.

Recent prior efforts have demonstrated the ability to identify MR with DNNs, though none have focused specifically on MVP. Vrudhula et al. previously trained a DNN model showing an ability to identify MR on echocardiogram(13), however did not examine performance in MVP patients making it unclear how it would perform in with MVP-related MR which has unique characteristics including being more often eccentric and late systolic dominant. Our results show that AI can achieve automated detection of MR even in patients with MVP, though the unique characteristics of MVP-related MR may explain our observed lower performance in this substratum. Long et al. (DELINEATE-MR) developed a fully automated end-to-end system that processes complete TTE studies, identifies color-Doppler clips, and performs ordinal MR grading using a large multi-center dataset(14). While their approach selects the three clips with the highest MR probability from any view, our design instead pre-specifies three clinically established mitral valve views (A2C, A4C, PLAX) to maintain anatomic consistency and alignment with guideline-based imaging for mitral valve disease. In contrast to their ordinal classification, we focused on the clinically actionable threshold of classifying ≥moderate-severe MR. Our multi-view architecture evaluates motion and jet characteristics across these standardized views, which may be of particular benefit to mitigate underestimation of eccentric MR commonly seen in MVP. Long et al. similarly reported decreased performance among MVP patients, corroborating our findings. When we manually reviewed echocardiographic videos of MVP patients which had been misclassified by the MR DNN, a highly eccentric MR jet resulting from MVP was the most common cause of misclassification, possibly explaining why the MR DNN had lower performance in MVP patients. If deployed in conjunction with the MVP DNN, the MR DNN would help to identify clinically significant MR in MVP patients, potentially leading to earlier surgical treatment and improved MVP patient outcomes.

Our work has several limitations to consider. Our external validation dataset from Houston Methodist DeBakey was limited to MVP cases only, which required us to construct a control dataset from UCSF echocardiograms to approximate external controls. While these non-MVP echocardiographic studies were drawn randomly from a very large potential population free of MVP likely approximating non-MVP studies from an external site, this was a hybrid external validation dataset with inherent limitations. We also lacked adjudication of MR in the external Houston Methodist dataset, limiting our ability to perform external validation for our MR DNN. Compared to other previously published DNNs for echocardiograms, our study had a relatively smaller sample size of MVP cases, owing in large part to the lower prevalence of this condition compared to other more common echocardiogram diagnoses. However, our UCSF MVP registry has the benefit of being confirmed MVP cases with all echocardiograms re-reviewed and accurately phenotyped by a cardiologist with expertise in MVP. Finally, the DNN models we present employed a unique multi-view architecture which simultaneously considers 3 echocardiographic views at once to make its assessment, whereas most other published DNNs only use 1 echocardiogram view. While our multi-view DNN architecture can outperform a single-view DNN in some settings, as we demonstrate here for MVP, it has the limitation of requiring the presence of all 3 views within a single echocardiogram study. In our case, this excluded 8 MVP patients out of over 700 patients. However, we also trained single-view DNNs as shown in our supplemental table, as these could be supplemented if necessary.

In conclusion, this study demonstrates that DNNs can achieve high performance for detecting MVP and clinically significant MR from echocardiographic videos. These algorithms could provide a novel approach for automated, accurate, and rapid diagnosis of MVP and its common clinical sequelae of MR across institutions. Further studies should evaluate the prospective and real-world deployment of these algorithms to examine their ability to enhance risk stratification and improve MVP related outcomes.

## Clinical perspectives

MVP DNN and MR DNN models have the potential to enhance follow-up care, guide imaging strategies, and support timely interventions, particularly in MVP patients at risk of major arrhythmic complications, cardiomyopathy and sudden cardiac death. Future work should include performance comparison with human experts to better evaluate the clinical utility of our model. Additionally, reader-in-the-loop studies will be valuable to assess whether our model can assist less experienced operators in improving their accuracy and consistency of MVP diagnosis.

## Data Availability

Data in this study was derived from clinical patient care and are not publicly available for privacy considerations.

## Supplementary tables

**Supplementary table 1:** Comparison of Single-view vs. Multi-view MVP models on UCSF test set.

|  | AUC | Sensitivity | Specificity |
| --- | --- | --- | --- |
| **A2C view model** | 0.862<br>(0.839-0.883) | 0.694<br>(0.644-0.743) | 0.858<br>(0.849-0.867) |
| **A4C view model** | 0.862<br>(0.840-0.881) | 0.739<br>(0.688-0.784) | 0.801<br>(0.791-0.812) |
| **PLAX view model** | 0.879<br>(0.860-0.898) | 0.752<br>(0.705-0.799) | 0.839<br>(0.828-0.849) |
| **Multi-view model** | 0.917<br>(0.899-0.934) | 0.797<br>(0.754-0.841) | 0.893<br>(0.884-0.901) |

## Conflicts of interest

GHT is an advisor to Viz.ai and Prolaio.

## Funding

This work was supported by the National Institutes of Health NHLBI R01HL153447 (Dr Delling and Dr Tison) and NIH DP2HL174046 (Dr. Tison). The sponsor had no role in study design, collection, management, analysis, interpretation of the data, and manuscript preparation and submission.

## Notes

### Author Declarations

Institutional Review Boards of the University of California San Francisco and Houston Methodist Hospital reviewed and provided ethical approval for this work.

